# Contrasting Evidence to Reimbursement Reality for Off-label use (OLU) of Drug Treatments in Cancer Care – Rationale and Design of the CEIT-OLU-project

**DOI:** 10.1101/19003152

**Authors:** AK Herbrand, AM Schmitt, M Briel, S Diem, H Ewald, A Hoogkamer, M Joerger, KA Mc Cord, U Novak, S Sricharoenchai, LG Hemkens, B Kasenda

**Affiliations:** Department of Medical Oncology, University Hospital Basel and University of Basel, Basel, Switzerland; Basel Institute for Clinical Epidemiology and Biostatistics, Department of Clinical Research, University Hospital Basel and University of Basel, Basel, Switzerland; Department of Health Research Methods, Evidence, and Impact, McMaster University, Hamilton, Canada; Department of Oncology and Hematology, Spital Grabs, Grabs, Switzerland; Department of Oncology and Hematology, Cantonal Hospital, St. Gallen, St. Gallen, Switzerland; Department of Immunobiology, Cantonal Hospital St. Gallen, St. Gallen, Switzerland; University Medical Library, University of Basel, Basel, Switzerland; Department of Medical Oncology, Bern University Hospital, Bern, Switzerland; Department of Hematology/Oncology and Palliative Care, Klinikum Stuttgart, Stuttgart, Germany

**Keywords:** Off-label use, evidence-based health care, reimbursement, cancer therapy

## Abstract

**Background:** Off-label drug use (OLU) reflects a perceived unmet medical need, which is common in oncology. Cancer drugs are often highly expensive and their reimbursement is a challenge for many health care systems. OLU is frequently regulated by reimbursement restrictions. For evidence-based health care, treatment ought to be reimbursed if there is sufficient clinical evidence for treatment benefit independently of patient factors not related to the treatment indication. However, little is known about the reality of OLU reimbursement and its association with the underlying clinical evidence. Here we aim to investigate the relationship of reimbursement decisions with the underlying clinical evidence.

**Methods/Design:** We extract patient characteristics and details on treatment and reimbursement of cancer drugs from over 3000 patients treated in three Swiss hospitals. We systematically search for clinical trial evidence on benefits associated with OLU in the most common indications. We will describe the prevalence of OLU in Switzerland and its reimbursement in cancer care, and use multivariable logistic regression techniques to investigate the association of approval/rejection of a reimbursement requests to the evidence on treatment effects and to further factors, including type of drug, molecular predictive markers and the health insurer.

**Discussion:** Our study will provide a systematic overview and assessment of OLU and its reimbursement reality in Switzerland. We may provide a better understanding of the access to cancer care that is regulated by health insurers and we hope to identify factors that determine the level of evidence-based cancer care in a highly diverse Western health care system.

## Background

Drugs prescribed in routine medical care typically require licensing and approval for defined indications by health authorities, for example the United States Food and Drug Administration (FDA), the European Medicines Agency (EMA), or Swissmedic (SM) in Switzerland. Indications usually contain restrictions in drug usage to a disease in a certain setting, line of treatment, and co-treatment with another drug. Off-label use (OLU) means using the drug outside the approved indication. In particular in cancer therapy, where new agents emerge frequently, OLU is common practice (1). International estimates range from 13% to 71% of adults receiving off-label therapy during cancer treatment (1), with about 30% in Switzerland (2). When new evidence for clinical benefit of a drug emerges, OLU allows usage before the indication is formally approved and labelled (3). One example is the human epidermal growth factor receptor 2 (HER2) directed agent trastuzumab, which is approved standard treatment for patients with HER2-positive breast cancer (4,5). Success of trastuzumab in HER2-positive breast cancer suggested benefits for other HER2-overexpressing or amplifying tumors as well, for example gastric cancer. Indeed, in 2010 a randomized controlled trial (RCT) showed better survival for patients with advanced gastric cancer overexpressing HER2 (6). However, it took another year until a formal approval for this new indication was granted by the FDA and in the meantime, patients could only receive this promising treatment via OLU. But counterexamples exist. Drugs being approved in certain cancer indications may not work in others, as shown for irinotecan and bevacizumab in colorectal cancer. Both drugs are effective and approved in the metastatic setting (7,8), however, in the adjuvant setting, no benefit was found (9,10); such example emphasizes that OLU may do harm to patients.

OLU is frequently regulated by reimbursement restrictions, because cancer drugs are often highly expensive and their reimbursement is a challenge for many health care systems. This may reduce costs, but to allow evidence-based health care, reimbursement should primarily be based on evidence. The Swiss healthcare system is highly diverse. Across the 26 cantons with diverse care structures and local regulation there are 57 statutory health insurers (as of Oct 2018) (11). They base their reimbursement policies on decisions of the Swiss Federal Office of Public Health (FOPH), which determines the drugs and indications that have to be reimbursed. For intended OLU, an upfront request for reimbursement to the patient’s health insurer is mandatory. These reimbursement requests are then appraised by reviewers (medical examiners) commissioned by the statutory health insurers, who give recommendations on acceptance of the request with subsequent access to that treatment. They may use various tools (12) to guide their decisions on whether to approve or disapprove the reimbursement for OLU and are supposed to consider the available clinical evidence but also the individual clinical circumstances and patient characteristics.

Although these tools are recommended, it is neither mandatory nor transparent how they would be applied. There is no structured and transparent systematic framework specifically regulating the case-by-case decisions. There is also no empirical information on the reimbursement reality, which would be a prerequisite for quality control and potential improvement of this approach. Overall, due to the high costs of oncology drugs, access of Swiss patients to drugs used in off-label indications is currently based on intransparent processes.

Here we describe the rationale and design of the OLU-Cancer project as part of the Comparative Effectiveness of Innovative Treatments (CEIT) project (13). The CEIT project evaluates the generation of clinical trial evidence for novel drug treatments, in the case of OLU using a large sample of patients to provide empirical evidence on access to OLU in a highly diverse Western health care system.

## Methods/Design

Our main objective is to investigate the relationship of reimbursement decisions to the underlying clinical evidence on patient benefits and subsequent access to cancer drugs. We also aim to assess the role of further factors that may influence the reimbursement. Finally, we aim to describe the prevalence of OLU and its reimbursement in cancer patients within the Swiss healthcare system.

The project is organized in three phases: First, we identify, extract and classify the data on OLU from patient records (electronic and paper-based). Second, we determine the clinical evidence using rapid review techniques, standard approaches for evidence synthesis, and review-of-review methods. Finally, we combine the results of both phases to address our research objectives.

### I Reimbursement reality

#### Eligibility Criteria

In August 2018, we started to screen patient records for reimbursement requests at three collaborating major hospitals, all being tertiary referral centers: University Hospital Basel (Department of Medical Oncology and Department of Hematology), University Hospital Berne (Department of Medical Oncology), and a large general district hospital in St. Gallen (Department of Oncology/Hematology).

We screen all patients who had their first consultation between January 2015 and July 2018. Patients were eligible if they have a malignant disease (solid or hematological) and receive systemic anti-cancer treatment with a drug or biologic. Patients with at least one reimbursement request issued by the treating physician will be included in data extraction (see figure 1). We exclude patients who have only a one-time appointment for a second opinion, whose disease is not treated with drugs but surgery or radiotherapy only, or for whom no reimbursement request was issued.

**Figure 1:**
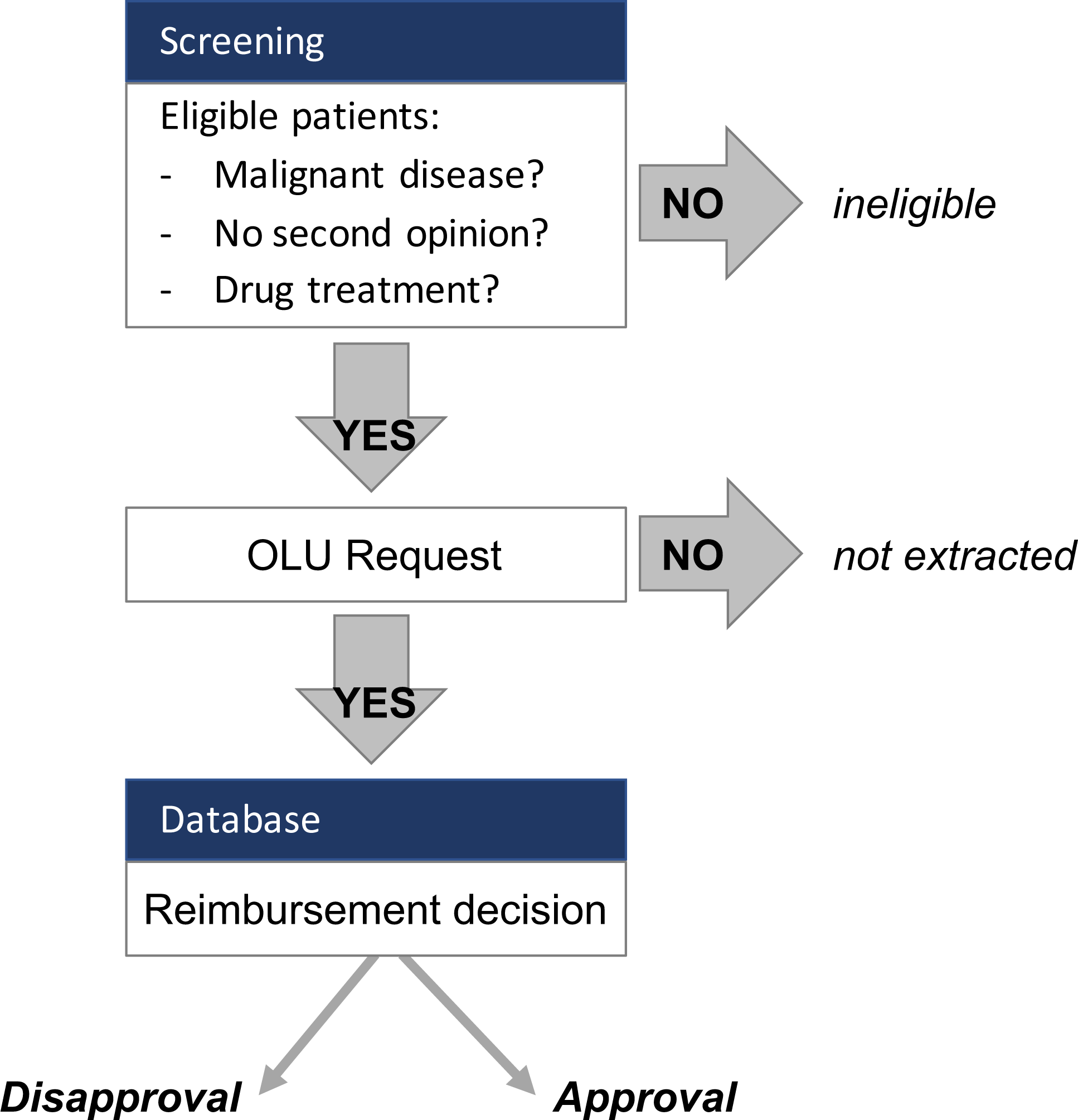
Flowchart for patient screening and eligibility criteria.

#### Sample size considerations

Based on a previous study on OLU in Switzerland (2), we assume that at least one reimbursement request for OLU has been issued for around 20% to 30% of eligible patients. In addition, preliminary data from the three sites revealed a proportion of 20% OLU. To account for the uncertainty on the actual frequency of OLU requests (both approved and disapproved), we consider a range of plausible proportions (15%, 20%, and 25%) of OLU among all patients. To achieve a high precision of estimates, we plan to screen around 3000 eligible patients. This which would result in the following frequencies and proportions with 95% confidence intervals (CIs) for OLU per patient:

- 450/3000 (15%, 95% CI: 13.8% to 16.3%)

- 600/3000 (20%, 95% CI: 18.6% to 21.5%)

- 900/3000 (30%, 95% CI: 28.4% to 31.7%)

We find all of them acceptable. To determine the actual proportion of disapproved reimbursement requests for OLU is a central objective of this project, however, based on preliminary data from this project and data from a single department that has implemented monitoring of its reimbursement requests since 2017, we assume that around 30% of OLU requests are disapproved. Considering the above-mentioned range of plausible frequencies of OLU requests, this would result in the following frequencies and proportions with 95% CIs for disapprovals per OLU request:

- 135/450 (30%, 95% CI 25.8% to 34.5%)

- 180/600 (30%, 95% CI 26.4% to 33.9%)

- 270/900 (30%, 95% CI 27.0% to 33.1%)

Thus, we consider a sample of 3000 eligible patients as sufficient to determine the prevalence of OLU, the prevalence of disapproved OLU, and also for our planned multivariable regression modelling (see III).

#### Extraction of patient-related health and reimbursement data

To assess the prevalence of OLU requests, we will screen patient records of all eligible patients for any reimbursement request. For those patients, we will extract patient demographics, disease and treatment details, and information on reimbursement requests and correspondence with the respective health insurer. This information will be stored in a relational database that we generated for this purpose. Details of the collected variables are available in the appendix (Table A1).

#### Definition and categorization of OLU

We will define OLU as any drug use that does not agree with the Swissmedic drug label. We will also assess unlicensed use (drug marketing is not licensed by Swissmedic for Switzerland, but approved by the FDA or EMA), and compassionate use (drug is neither licensed by the FDA, EMA nor Swissmedic) (see table 1). We will further sub-categorize OLU according to disease, intended treatment setting (e.g. adjuvant), line of treatment (e.g. first-line) and use as single agent or drug combination. This allows us to investigate the prevalence of OLU in different situations.

**Table 1:**
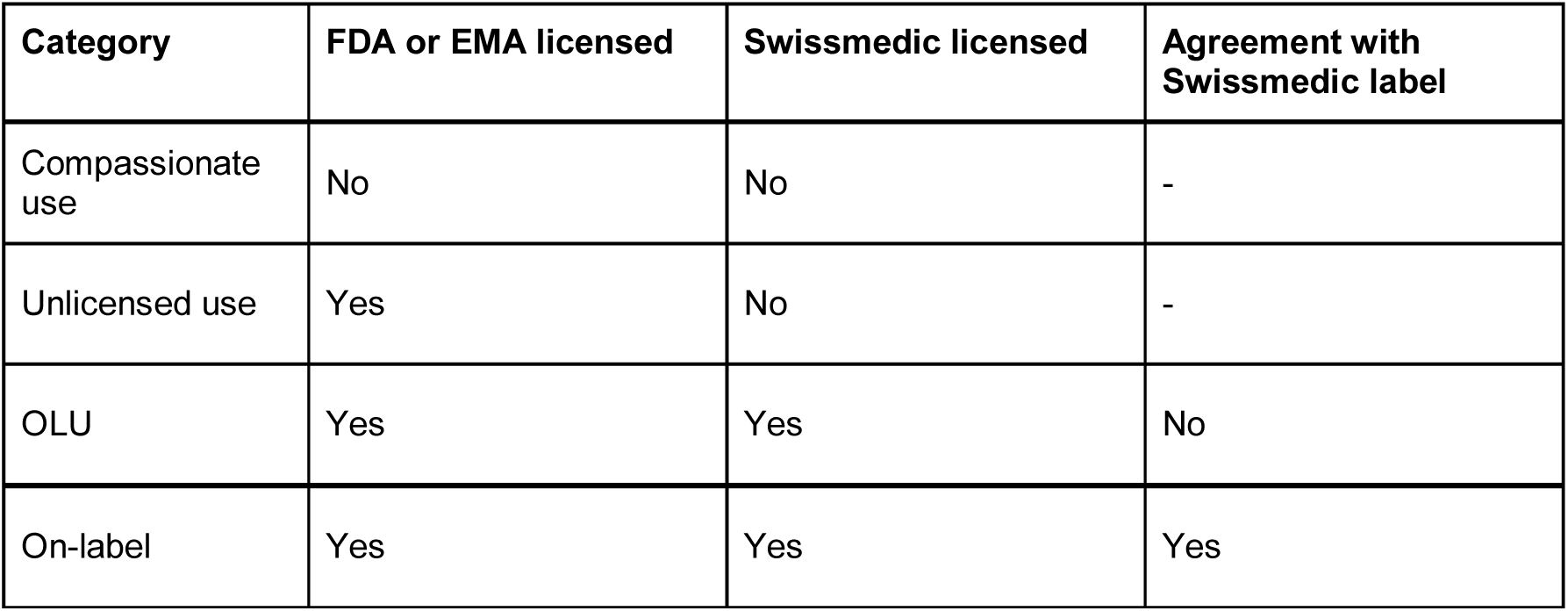
Categorization of drug use depending on license, label and limitations. Abbr.: FDA= Food and Drug Association, EMA= European Medicines Agency

| Category | FDA or EMA licensed | Swissmedic licensed | Agreement with Swissmedic label |
| --- | --- | --- | --- |
| Compassionate use | No | No | - |
| Unlicensed use | Yes | No | - |
| OLU | Yes | Yes | No |
| On-label | Yes | Yes | Yes |

#### Timeline of licensing and approval

Licensing of a cancer drug often starts with a single indication and, over time, additional indications are approved. Drug labels and on- and off-label status may change over time. We will systematically record the approval dates of all requested drugs for each disease and new indication in a structured database. We will identify these dates by searching for publications and press releases of Swissmedic and the FOPH to develop a timeline for each drug and its label changes.

Two trained oncologists/hematologists will independently determine the OLU status for each request at the time of the request. Disagreements will be solved by discussion. Because the off-label status is time-dependent, we will match the date of the OLU request with the approval-timeline of the respective drug label.

#### OLU prevalence

We will estimate the prevalence of OLU and the frequency of reimbursement. Specifically, we will investigate:

- The frequency and proportion of all reimbursement requests among all eligible patients treated for a malignant disease.
  - The frequencies and proportions of OLU, compassionate and unlicensed use among all reimbursement requests.
  - Frequency and proportion of OLU requests for a drug targeting a molecular predictive marker irrespective of tumor site of origin.
- Frequency and proportion of approval/rejection among all reimbursement requests for OLU, unlicensed and compassionate use.
  - Frequency and proportion of how costs for requested drugs are covered stratified by approval/rejection
  - Frequency and proportion of rebuttals to rejections among all reimbursement requests for OLU, unlicensed and compassionate use
  - Frequency of how many decisions were changed from rejection to approval by rebuttal of the treating physician
- The time between reimbursement request and approval/disapproval of reimbursement decision. Unit of calculation is the individual request. We will re-evaluate all these factors in subgroups of requests clustered by the patient’s health insurer (insurer as per index date of first reimbursement request).

### II Clinical evidence

We will systematically evaluate the evidence for treatment effects on overall survival (OS) and progression-free survival (PFS) in OLU situations.

#### Literature search

For the most common off-label indications, we will generate overarching clinical questions following the PICO structure (14), composed of population (disease, treatment setting, treatment line), intervention (drug, single agent or combination), control (any comparator), and outcome (PFS or OS). Most common OLU indications and resulting PICO questions will be derived from the extracted patient data (for details see appendix, table A1).

We will use rapid review techniques to identify systematic reviews (SRs) of RCTs addressing these questions. If there are no eligible systematic reviews available, we will directly search for RCTs. We will search PubMed and use pertinent keywords and subject headings for cancer sites and generic drug terms. Standard search filters for SRs and RCTs will be used (15). In general, we will include publications reporting treatment effects on PFS and OS that are published in English or German. No further limits will be applied. Further evidence will be sought by backward citation chasing (16) (screening the reference lists of included publications). Search strategies will be created by an information specialist and will be peer-reviewed.

We will additionally evaluate literature provided by the requesting physician and consult guidelines provided by the European Society of Medical Oncology (ESMO) and verify if our search identifies any RCTs not found in our systematic literature searches (17).

#### Study selection and data extraction

Overall, for feasibility and efficiency reasons, we use the SRs primarily to identify pertinent RCTs and their results. In a first step, we will select the most recent systematic review publication identified in the SR search that is covering the PICO. We will always prefer a recent Cochrane Review due to the comparatively high methodological quality and standardized and transparent reporting. We only consider SRs that are sufficiently transparent in reporting key methods. This means, we include only SRs that adequately report the PRISMA items “identification”, “screening”, “eligibility”, “included” (18). We deem reporting adequate if it would let us replicate the search for RCTs. Selection of SRs will be done independently by two reviewers and confirmed by a third reviewer in case of uncertainty.

In a second step, we will identify the pertinent RCTs included in eligible SRs. From each eligible SR article, we extract bibliographic details of the SR, search details (search date, databases), and any RCT and corresponding publication matching our PICO of interest. From each pertinent RCT article, we extract bibliographic information and the date of publication (earliest available: online or printed if printed publication was available before published online). We will extract the treatment effects directly from the RCT publication, based on the main analysis of the intention to treat population (ITT), where available. The rationale behind this is that we want to depict the evidence as it was at the time of request. In case of multiple reported effect estimates, we prefer that one clearly labeled as prespecified, main analysis, or first reported in the abstract (acknowledging possible spin by relying on prominently reported results for consistency, which will be addressed in sensitivity analyses).

If no appropriate SR can be identified, we will apply the same procedure to the RCT search.

#### Evidence synthesis and overview

We will synthesize the evidence by using cumulative meta-analysis techniques (19,20). We will combine all trials on the same PICO question and determine for the time point, when new evidence is available, the current knowledge on clinical benefits (that is the summary treatment effect estimate for OS and PFS from all RCTs up to that point). This allows us to describe the evolution of clinical knowledge from RCTs in any oncological off-label indication with clinical trial evidence over time.

We will categorize the available evidence on patient-relevant treatment benefits from RCTs (summary treatment effect from meta-analyses) for each drug and indication into:

1. The combined analysis shows OS benefit (nominal statistical significance).
2. The combined analysis shows PFS benefit (nominal statistical significance), but no OS benefit.
3. No indication for benefit or harm (no nominal statistically significant OS or PFS effect).
4. Inconclusive (nominal statistical treatment benefit for OS but significant harm for PFS or vice versa).
5. The combined analysis shows OS harm.
6. The combined analysis shows PFS harm.
7. Unclear (no randomized trial evidence available).

### III Contrasting Evidence to Reimbursement Reality

Finally, we will investigate the association between the reimbursement decision for OLU and clinical evidence using multilevel multivariable logistic regression. Furthermore, we aim to identify other factors than clinical evidence explaining any potential discrepancies between clinical evidence and reimbursement.

#### Association of reimbursement to evidence

We will use multilevel multivariable logistic regression to investigate the association between the clinical evidence supporting OLU and the likelihood for reimbursement of OLU. We have chosen multilevel modelling, because requests are clustered by patients and health insurers. We will condense the seven categories of clinical evidence into three groups (A-C) and summarize treatment benefits:

A. Clear patient-relevant benefit (corresponds to category 1)
B. Potential patient-relevant benefit (corresponds to category 2)
C. No proven patient-relevant benefit (corresponds to category 3-7)

We will determine the category for the time when each reimbursement was requested. Category A will be the reference for this categorical variable, which is the independent variable in the regression analysis. Rejection of a request for reimbursement of OLU will be the dependent binary variable (rejected versus approved). The association will be expressed as odds ratios (OR) with 95% confidence intervals, with an OR>1 indicating a higher likelihood for the reimbursement to be rejected by the health insurer. We hypothesize that the risk for rejection will be higher in situations of category C than B and higher for category B than A.

The above-mentioned consideration of time points in our data collection and analysis will allow us to investigate potential time lags between occurrence of new evidence for treatment benefit and Swissmedic approval/labeling. In such periods, patients can only access new drugs through OLU reimbursement by the health insurer or extended access programs by the respective drug manufacturer. However, the latter is completely at the discretion of the drug manufacturer, therefore, requesting OLU from the health insurer is often the only way to receive the drug. We will therefore additionally investigate the following:

- The time lag between availability of evidence for treatment benefit (category A or B) and approval of the drug by Swissmedic. This time will be calculated from the earliest date of trial publication to the date of approval by Swissmedic.
- The number of cases in this time period, for each indication, in which an OLU request was issued and the number of these requests rejected for reimbursement, although evidence for treatment benefit (category A or B) was available.

#### Association of reimbursement to other factors

To investigate whether patient or disease characteristics that are not related to the treatment indication influence the risk for rejection of request for OLU reimbursement, we will perform multilevel multivariable logistic regression analysis including seven variables. These variables are:

1. Sex (categorical variable: male vs. female)
2. Age (continuous variable)
3. Presence of molecular predictive marker (binary variable; we anticipate that presence of a marker lowers the risk of rejection because of the availability of a drug target independent of the disease)
4. Disease type (categorical variable: solid vs. hematological malignancies; we anticipate that chances for reimbursement are higher for hematological malignancies because there are often only OLU treatment options available for hematological malignancies)
5. Type of treatment (categorical variable: immunotherapy [checkpoint inhibition] vs. all others; we anticipate that chances for reimbursement are higher for immunotherapy because this drug class has recently been approved for several tumor entities)
6. Orphan drug status (binary variable; we anticipate that chances for reimbursement are higher for an orphan drug/indication because there are less treatment options available)
7. Tumor incidence (continuous variable; we anticipate that chances for reimbursement are higher for rare diseases because there are less treatment options available)

We assume that there is less evidence for rare diseases. However, such patients may often be particularly in need for OLU. We will rank the entities by incidence according to global cancer statistics as issued by the WHO (21) and label orphan drugs according to Swissmedic (22). For sensitivity analyses, we will consider the orphan drug designation as used by the FDA (23).

### Handling of missing data

We will report frequencies (proportions) of missing data of all independent and dependent variables. The primary analysis will be based on complete cases, but we will use multiple imputations for missing data for a sensitivity analysis using all cases (24).

### Software

We will use Ninox (https://ninoxdb.de/de/) for managing data and generating relational databases, R for data cleaning and most statistical analyses (The R Project for Statistical Computing, https://www.r-project.org/), and Stata (StataCorp. 2017. Stata Statistical Software: Release 15. College Station, TX: StataCorp LLC) for multilevel regression modelling.

## Discussion

This study will provide a comprehensive assessment of OLU and its reimbursement reality in Switzerland, a systematic overview of the comparative effectiveness of OLU in cancer care, and a thorough investigation on the association between OLU reimbursement and the underlying evidence from clinical research. By publishing this rationale and design paper, we intend to minimize ambiguity and increase transparency concerning our study (25).

To our knowledge, this will be the first study targeting OLU in such extent and level of detail. We will use transparent sources and apply systematic and reproducible search strategies. The collaboration with three hematology/oncology departments across Switzerland will provide us with a sufficient number of patients to gain a representative sample for our planned investigations.

We will highly depend on the comprehensiveness of documentation in patient files, but we expect that the documentation of correspondence with the health insurer will at least be routinely included in the patient files due to the substantial economic and administrative impact.

This study will provide a comprehensive overview on access to off-label treatments in a highly developed country, characterized by a fragmented and expensive health care system. It will help to determine if access to perceived unmet medical needs is based on clinical evidence and if the Swiss health care system employs principles of evidence-based health care for OLU. We hope to reveal potential inequalities in access, to better understand and ultimately improve the quality of reimbursement decisions for cancer patients.

## Data Availability

Not applicable.

## Abbreviations

CEIT: Comparative Effectiveness of Innovative Treatments
EMA: European Medicines Agency
FDA: Food and drugs association
FOPH (Swiss): Federal Office of Public Health
HER2: Human epidermal growth factor receptor 2
OLU: Off-label Use
OS: Overall survival
PFS: Progression free survival
PICO: Population, intervention, control, outcome
RCT: Randomized controlled study
SM: Swissmedic
SR: Systematic review

## Declarations

### Ethics approval and consent to participate

We will use routinely collected health data of patients who did not explicitly give informed consent to our study. The responsible research ethics committee reviewed this project and approval was granted (EKNZ Project ID 2018-01431, approved 10th of August 2018).

All study personnel with access to individual patient data will sign confidentiality agreements. Patient data will be handled using a specific privacy securing system.

### Consent for publication

Not applicable.

### Availability of data and material

Not applicable.

### Authors’ contributions

Drafting of the manuscript: AKH, AMS, LGH, BK. Critical revision of the manuscript for important intellectual content: MB, HE, SD, UN, MJ. Collection of data: AH, KAM, SS. All authors read and approved the final manuscript.

## Acknowledgements

Not applicable.

## References

1. Saiyed MM, Ong PS, Chew L. Off□label drug use in oncology: a systematic review of literature. J Clin Pharm Ther [Internet]. 2017; Available from: https://onlinelibrary.wiley.com/doi/abs/10.1111/jcpt.12507

2. Joerger M, Schaer-Thuer C, Koeberle D, Matter-Walstra K, Gibbons-Marsico J, Diem S, et al. Off-label use of anticancer drugs in eastern Switzerland: a population-based prospective cohort study. Eur J Clin Pharmacol. 2014 Jun;70(6):719–25.

3. Stafford RS. Regulating off-label drug use--rethinking the role of the FDA. N Engl J Med. 2008 Apr 3;358(14):1427–9.

4. Senkus E, Kyriakides S, Penault-Llorca F, Poortmans P, Thompson A, Zackrisson S, et al. Primary breast cancer: ESMO Clinical Practice Guidelines for diagnosis, treatment and follow-up. Ann Oncol. 2013 Oct;24 Suppl 6:vi7–23.

5. Cardoso F, Costa A, Norton L, Senkus E, Aapro M, André F, et al. ESO-ESMO 2nd international consensus guidelines for advanced breast cancer (ABC2)†. Ann Oncol. 2014 Oct;25(10):1871–88.

6. Van Cutsem E, Kang Y, Chung H, Shen L, Sawaki A, Lordick F, et al. Efficacy results from the ToGA trial: a phase III study of trastuzumab added to standard chemotherapy in first-line HER2-positive advanced gastric cancer. J Clin Oncol. 2009;27(18)(suppl):LBA4509.

7. Douillard JY, Cunningham D, Roth AD, Navarro M, James RD, Karasek P, et al. Irinotecan combined with fluorouracil compared with fluorouracil alone as first-line treatment for metastatic colorectal cancer: a multicentre randomised trial. Lancet. 2000 Mar 25;355(9209):1041–7.

8. Hurwitz H, Fehrenbacher L, Novotny W, Cartwright T, Hainsworth J, Heim W, et al. Bevacizumab plus irinotecan, fluorouracil, and leucovorin for metastatic colorectal cancer. N Engl J Med. 2004 Jun 3;350(23):2335–42.

9. Saltz LB, Niedzwiecki D, Hollis D, Goldberg RM. Irinotecan fluorouracil plus leucovorin is not superior to fluorouracil plus leucovorin alone as adjuvant treatment for stage III colon cancer: results of CALGB …. Journal of Clinical [Internet]. 2007; Available from: http://citeseerx.ist.psu.edu/viewdoc/download?doi=10.1.1.977.2822&rep=rep1&type=pdf

10. de Gramont A, Van Cutsem E, Schmoll H-J, Tabernero J, Clarke S, Moore MJ, et al. Bevacizumab plus oxaliplatin-based chemotherapy as adjuvant treatment for colon cancer (AVANT): a phase 3 randomised controlled trial. Lancet Oncol. 2012 Dec;13(12):1225–33.

11. Bundesamt für Gesundheit. Verzeichnisse der zugelassenen Kranken-und Rückversicherer [Internet]. [cited 2019 May 1]. Available from: https://www.bag.admin.ch/bag/de/home/versicherungen/krankenversicherung/krankenve rsicherung-versicherer-aufsicht/verzeichnisse-krankenundrueckversicherer.html

12. von Stokar T, Vettori A, Fliedner J. Zugangsgerechtigkeit und -sicherheit bei Krebsmedikamenten im Off-label Use. Krebsliga Schweiz (KLS), Bern [Internet]. 2013; Available from: https://assets.krebsliga.ch/downloads/130527_bericht_zusammenfassung_off_label_use_infras_d.pdf

13. Ladanie A, Speich B, Naudet F, Agarwal A, Pereira TV, Sclafani F, et al. The Comparative Effectiveness of Innovative Treatments for Cancer (CEIT-Cancer) project: Rationale and design of the database and the collection of evidence available at approval of novel drugs. Trials. 2018 Sep 19;19(1):505.

14. Richardson WS, Wilson MC, Nishikawa J, Hayward RS. The well-built clinical question: a key to evidence-based decisions. ACP J Club. 1995 Nov;123(3):A12–3.

15. Lefebvre C, Manheimer E, Glanvill J. Chapter 6: Searching for Studies. In: Higgins JPT & Green S (editors). Cochrane Handbook for Systematic Reviews of Interventions: Version 5.1.0 (updated March 2011). The Cochrane Collaboration, 2011. Available from: http://handbook-5-1.cochrane.org/

16. Cooper C, Booth A, Britten N, Garside R. A comparison of results of empirical studies of supplementary search techniques and recommendations in review methodology handbooks: a methodological review. Syst Rev. 2017 Nov 28;6(1):234.

17. ESMO guidelines [Internet]. Available from: https://www.esmo.org/Guidelines/ESMO-MCBS

18. Moher D, Liberati A, Tetzlaff J, Altman DG, PRISMA Group. Preferred reporting items for systematic reviews and meta-analyses: the PRISMA statement. BMJ. 2009 Jul 21;339:b2535.

19. Sutton AJ, Higgins JPT. Recent developments in meta-analysis. Stat Med. 2008;27(5):625–50.

20. Lau J, Schmid CH, Chalmers TC. Cumulative meta-analysis of clinical trials builds evidence for exemplary medical care. J Clin Epidemiol. 1995 Jan;48(1):45–57; discussion 59–60.

21. Fact Sheets by Population [Internet]. Available from: http://globocan.iarc.fr/Pages/fact_sheets_population.aspx

22. Swissmedic. Orphan drug status. [Internet]. 2018. Available from: https://www.swissmedic.ch/swissmedic/de/home/ueber-uns/swissmedic--schweizerisches-heilmittelinstitut/patienten-und-anwender.html

23. US Food and Drug Administration. Search orphan drug designations and approvals. [Internet]. 2014; Available from: https://www.accessdata.fda.gov/scripts/opdlisting/oopd/

24. Sterne JAC, White IR, Carlin JB, Spratt M, Royston P, Kenward MG, et al. Multiple imputation for missing data in epidemiological and clinical research: potential and pitfalls. BMJ. 2009 Jun 29;338:b2393.

25. Godlee F. Publishing study protocols: Making them visible will improve registration, reporting and recruitment. Vol. 2. 2001.

26. Spezialitätenliste (SL) - Übersicht [Internet]. [cited 2018 Jul 12]. Available from: http://www.xn--spezialittenliste-yqb.ch/

